# Estimates of regional infectivity of COVID-19 in the United Kingdom following imposition of social distancing measures

**DOI:** 10.1101/2020.04.13.20062760

**Authors:** Robert Challen, Krasimira Tsaneva-Atanasova, Martin Pitt, Tom Edwards, Luke Gompels, Lucas Lacasa, Ellen Brooks-Pollock, Leon Danon

## Abstract

The SARS-CoV-2 reproduction number has become an essential parameter for monitoring disease transmission across settings and guiding interventions. The UK published weekly estimates of the reproduction number in the UK starting in May 2020 which are formed from multiple independent estimates. In this paper, we describe methods used to estimate the time-varying SARS-CoV-2 reproduction number for the UK. We used multiple data sources and estimated a serial interval distribution from published studies. We describe regional variability and how estimates evolved during the early phases of the outbreak, until the relaxing of social distancing measures began to be introduced in early July. Our analysis is able to guide localised control and provides a longitudinal example of applying these methods over long timescales.

## Introduction

In late 2019 an outbreak of a novel infectious disease was detected. It manifested principally with severe acute respiratory distress, and pneumonia, [1] although many cases followed a mild course [2]. The pathogen was rapidly identified as a new species of coronavirus (severe acute respiratory syndrome coronavirus 2, SARS-CoV-2), and the disease named COVID-19 [3]. Global transmission of the virus followed and major outbreaks have been observed in Europe, beginning with Italy [4]. On the 31st Jan 2020 the first cases were identified in the UK [5]. This was initially managed using testing of suspected individuals in the community, contact tracing and isolation of affected cases. However this was successful only in delaying the spread of the disease and on 13th March 2020 the UK government moved towards a mitigation strategy reserving testing for hospital inpatients only [6]. Following this, a step wise implementation of social distancing measures were mandated by the government including voluntary self isolation of any symptoms & vulnerable people [7], a ban on non essential travel worldwide [8] and school closures [9]. Finally on 23rd March 2020 the government mandated that everyone apart from essential workers should stay at home and away from others [10], instituting a countrywide “lock-down”.

Epidemiological studies conducted during the outbreak in China have provided us with a number of estimates of the parameters describing the virus’s spread through the population including a reproduction number between 2.24 and 3.58 [11] and a median incubation period of 5.1 days (credible interval 4.5 to 5.8) [12]. It is estimated that fewer than 2.5% of people will show signs before 2.2 days and 97.5% of people who will develop symptoms will have done so by 11.2 days after exposure [12].

We investigated the reproduction number of SARS-CoV-2 in the UK to determine whether there are any spatial or temporal patterns beyond those resulting from the imposition of social distancing measures, and to track the progression of the outbreak. This article summarizes the methodology and interpretation of national and regional estimates of the time varying reproduction number (*R*_*t*_) of the SARS-CoV-2 outbreak in the United Kingdom. These estimates were provided to the Scientific Pandemic Influenza Group on Modelling (SPI-M) [13] and formed part of the weekly UK *R*_*t*_ estimates [14].

## Methods

This section describes the data sources, their processing and combination and our methodology for estimating the serial interval of SARS-CoV-2 infections, and calculation of *R*_*t*_ estimates.

### Data

We integrate data from a variety of sources, both publicly available and provided to the Scientific Pandemic Influenza Group - Modelling (SPI-M) [13] by Public Health England (PHE) and the Defence Science and Technology Laboratory (DSTL) [15]. We use these data to estimate *R*_*t*_ and the exponential growth rate for the UK as a whole, for the four nations of the UK (CTRY), and for the seven NHS regions in England (NHSER).

At the UK level, data are available on cases and deaths through the PHE coronavirus tracker [16]. The tracker publishes an overall number of cases and deaths in the UK on a daily basis. It also provides a regional breakdown of the 4 nations, which exclude tests performed in private laboratories (Pillar 2 tests). At the time of this analysis, historical time series of the UK level data was not made available though the PHE site, however this information was collected prospectively and curated by Tom White’s aggregated COVID 19 UK data github site [17]. Cumulative case counts from both the PHE headline UK figure and the combined sum of the 4 nations are compared as these numbers differ.

Hospital admission data are available from NHS trusts across the UK via the DSTL, which in turn aggregate the situation reports (SitReps) provided by NHS hospital trusts. These are aggregated to UK level. Although referred to here as “hospital admission” it includes admissions of patients who are subsequently identified as COVID-19 cases in hospital, but identified in hospital, and patients with known COVID-19 who are then admitted to hospital.

For the four devolved nations of the UK we used different data sources for each nation. In England we used data from the SPI-M provided line lists for cases and deaths in England. Cases are restricted to those processed in NHS labs (Pillar 1), and are available by date of specimen collection. For the other three nation states of the UK, as above, both historical cases and death data are aggregated from Public Health Wales [18] and Scotland’s [19] sites, and from Northern Ireland’s HSC site [20] respectively, using a time series retrieved from Tom White’s aggregated COVID 19 UK data github site [17] for Scotland, Wales and Northern Ireland. Death data from the 4 nations are provided via the CHESS data set [21]. Death data are subject to a weekly periodicity due to reporting delay over the weekend which is mitigated by using date of death rather than date of report.

At the level of the NHS England regions, we take the case data from anonymised test result line listings. Mortality data for NHS England is provided by PHE in a canonical line list of deaths, and is used alongside the CHESS data set[21]. Admissions data is available from DSTL feeds, and ICU admissions from CHESS data set[21].

For England as a nation and for the NHS England regions we also have data available from triage telephone calls from NHS 111 and 999 services. The data on the calls made to 111 & 999 as well as the outcome of that call are provided as aggregate numbers broken down by age.

For all data sources cumulative case figures are converted into daily incidence figures, and any data which is broken down by age, or by gender, are combined. In case and death data, the final 5 days of the time series are discarded to account for possible reporting delay. The resulting time series are analysed for outlying data points, which are more than 5 standard deviations away from the mean of the nearest 14 data points. Outlying or missing data are imputed from a linear interpolation of the logarithm of incidence figures, implemented in the R ‘forecast’ library [22], and the results are truncated to ensure no negative incidence figures. A smoothing function (a linear spline interpolation) is then fitted to the logarithm of incidence applied over a 7 day window [23]. This is needed as all the data sources have some degree of weekly periodicity regardless of source.

### Combination of data sources

Our estimates are based on an aggregation of the various data sources described above. We generate four estimates of *R*_*t*_ based on single time series from each data stream:

a. deaths (EpiEstim/Deaths),
b. cases (EpiEstim/4NationsCases),
c. telephone triage (EpiEstim/Triage),
d. hospital admissions.

The aggregation procedure is kept simple and combines multiple data sources when applicable using the mean. Any possible biases this introduces are consistent throughout the time series so that it does not affect relative changes and hence either estimates of *R*_*t*_ or growth rates. The resulting time series are manually inspected for consistency and to check there are not abrupt changes in the data streams. The source of the combined data sets and more information about the processing steps used is shown in Supplemental table 1.

**Table 1:** Estimates of the value of *R*_*t*_ in the UK on 4th July 2020

| Observation | $R(t)$ (95% CI) | Count per 1M per day (95% CI) |
| --- | --- | --- |
| Cases | 0.87 (0.81; 0.92) | 2.4 (2.0; 2.9) |
| Deaths | 0.91 (0.84; 0.99) | 1.4 (1.3; 1.5) |
| Admissions | 0.81 (0.76; 0.86) | 2.2 (2.0; 2.4) |

### The serial interval of SARS-CoV-2 infections, *R*_*t*_, and time delay of estimates

The serial interval and the generation interval are closely related measures. The generation interval is a measure of the time taken for an infection to pass from one person (infector) to another (infectee) in a chain of transmission. The serial interval, on the other hand, is a measure of the time between the appearance of clinical symptoms in the infector and infectee. The generation interval cannot be observed directly as both infection events are only detectable once the virus has incubated and become symptomatic. The serial interval is often used as an observable proxy for the generation interval. The use of the serial interval as a proxy for the generation interval is known to produce biased estimates for the reproduction number [24,25], but has the advantage of being directly observed through contact tracing.

Assumptions about the serial interval of SARS-CoV-2 have an impact on the absolute level of our estimates of *R*_*t*_. We have used multiple approaches to estimate the serial interval. Firs, a UK specific serial interval was calculated from early case tracking data (the FF100 case data provided by DSTL). Second, a literature review was conducted and the serial intervals from a range of sources [4,26–35] pooled [36]. The serial interval can be well described by a truncated empirical distribution, with a mean plus 95% credible interval of 5.59 days (5.09; 6.20), and a standard deviation of 4.15 days (3.94; 4.46). This uncertainty in the serial interval distribution is propagated to our estimates of *R*_*t*_ and used to determine confidence intervals. In other work we have analysed the effect of the pragmatic use of serial interval instead of generation interval and find the effect on estimates of Rt to be small (∼5% of the absolute value) when it is close to one. This choice does not influence the estimate of time when Rt transitions from growth to decay [36].

Using the inferred serial interval distribution, we analysed the time series data using forward equation method [37], implemented in the EpiEstim R library [37–39], to estimate the time varying reproduction numbers during the outbreak.

The renewal equation method is predicated on a time series of infections, and on the infectivity profile - a measure of the probability that a secondary infection occurred on a specific day after the primary case, given a secondary infection occurred [37]. A Bayesian framework is then used to update a prior probabilistic estimate of *R*_*t*_ on any given day with both information gained from the time series of infections in the epidemic to date and the infectivity profile to produce a posterior estimate. In both the original [38] and revised [39] implementations of this method, the authors acknowledge the pragmatic use of the serial interval distribution, as a proxy measure for the infectivity profile, and the incidence of symptom onset or case identification as a proxy for the incidence of infection, with the caveat that these introduce a time lag into the estimates of *R*_*t*_. We make the simplifying assumption that there is negligible mixing of populations between each geographical area and treat each location independently.

The method uses a sliding time window during which the instantaneous reproduction number is assumed to be constant. We used both a 7 day and a 28 day sliding window for calculations of the time varying reproduction number which provides two estimates with alternative trade offs between noise and loss of detail. Our *R*_*t*_ estimate is calculated using a loosely informed prior estimate of *R*_*t*_ as a gamma distribution with mean of 1 and standard deviation 2. This prior distribution is based on an assumption of the approximate value of *R*_*t*_ on the conditions following lock-down, rather than reflecting values of *R*_*0*_ commonly described in the literature [11] as those are based on the situation without social distancing measures in place.

Other events in the timeline of an infection also serve as a proxy for observations of infections in the past including positive testing, hospitalization for severe disease, or death. Although best practice is to calculate *R*_*t*_ using a generation interval distribution and infection events [25], neither the generation interval distribution nor the infection event data can be directly observed, and inferring them can also introduce potential bias and uncertainty. As a pragmatic initial step, we use the serial interval distribution described above in lieu of the generation interval, and the various observations available to us, including triage contacts, cases, admissions and deaths as a proxy of prior transmission events, and compare those results.

There is a time delay between infection, symptom onset, case identification, hospital admission and ultimately death, and this affects the timing of our estimation of *R*_*t*_. In a separate analysis we estimated these time delays (shown in Supplemental table 2) and apply these estimates as a correction to the time of our estimates of *R*_*t*_ to align them to the date of presumed infection.

**Table 2:** Estimates of Mean *R*_*t*_ and 95% confidence intervals for the individual countries in the United Kingdom, based on cases, deaths and hospital admissions on the 4th July 2020

| Country | Observation | R(t) (95% CI) | Count per 1M per day (95% CI) |
| --- | --- | --- | --- |
| England | Cases | 0.85 (0.83; 0.88) | 2.4 (2.0; 2.8) |
|  | Deaths | 0.90 (0.86; 0.93) | 1.6 (1.5; 1.7) |
|  | Admissions | 0.84 (0.81; 0.86) | 2.5 (2.3; 2.7) |
|  | Triage | 0.94 (0.93; 0.96) | 14.6 (13.2; 16.1) |
| Northern Ireland | Deaths | 0.70 (0.31; 1.32) | 0.2 (-0.0; 0.4) |
|  | Admissions | 0.72 (0.44; 1.09) | 0.4 (0.2; 0.7) |
| Scotland | Cases | 0.83 (0.75; 0.93) | 1.3 (0.9; 1.8) |
|  | Admissions | 0.74 (0.53; 1.00) | 0.1 (0.0; 0.2) |
| Wales | Cases | 0.85 (0.81; 0.90) | 6.5 (4.7; 8.8) |
|  | Deaths | 0.79 (0.63; 0.99) | 1.0 (0.7; 1.3) |
|  | Admissions | 0.87 (0.76; 0.98) | 1.7 (1.4; 2.2) |

Code and processed data involved in this analysis are available on GitHub [40].

## Results

### UK Overview

Our estimate of the median value of *R*_*t*_ based for the United Kingdom based on cases, deaths and hospital admissions from the 4th July 2020 is presented in Table 1.

In Figure 1A, we show the incidence per million people of cases, deaths, and hospital admissions due to COVID-19 in the UK over the outbreak. Panel B shows the associated values for *R*_*t*_. The 3 data sources show similar patterns of exponential increase, with admissions and deaths lagging cases followed by a slower phase of exponential decline, beginning 2-3 weeks after the lock-down. In Figure 1B we see the estimates of *R*_*t*_ corrected to date of infection. There is a prominent single initial peak in *R*_*t*_ from admissions and deaths in late February, followed by a decline over the course of the next few months. *R*_*t*_ crosses 1 at the beginning of April, after which it remains below 1 during the lock-down period. Estimates based on cases show a biphasic pattern with an initial peak in mid February and a second smaller peak in early March, follow the same pattern as other estimates. The peak values of *R*_*t*_ vary by observation with peak *R*_*t*_ by admissions being 8.3, by cases 4.8 and by deaths 3.3. These are within the confidence limits of estimates described in other countries [11].

**Figure 1:**
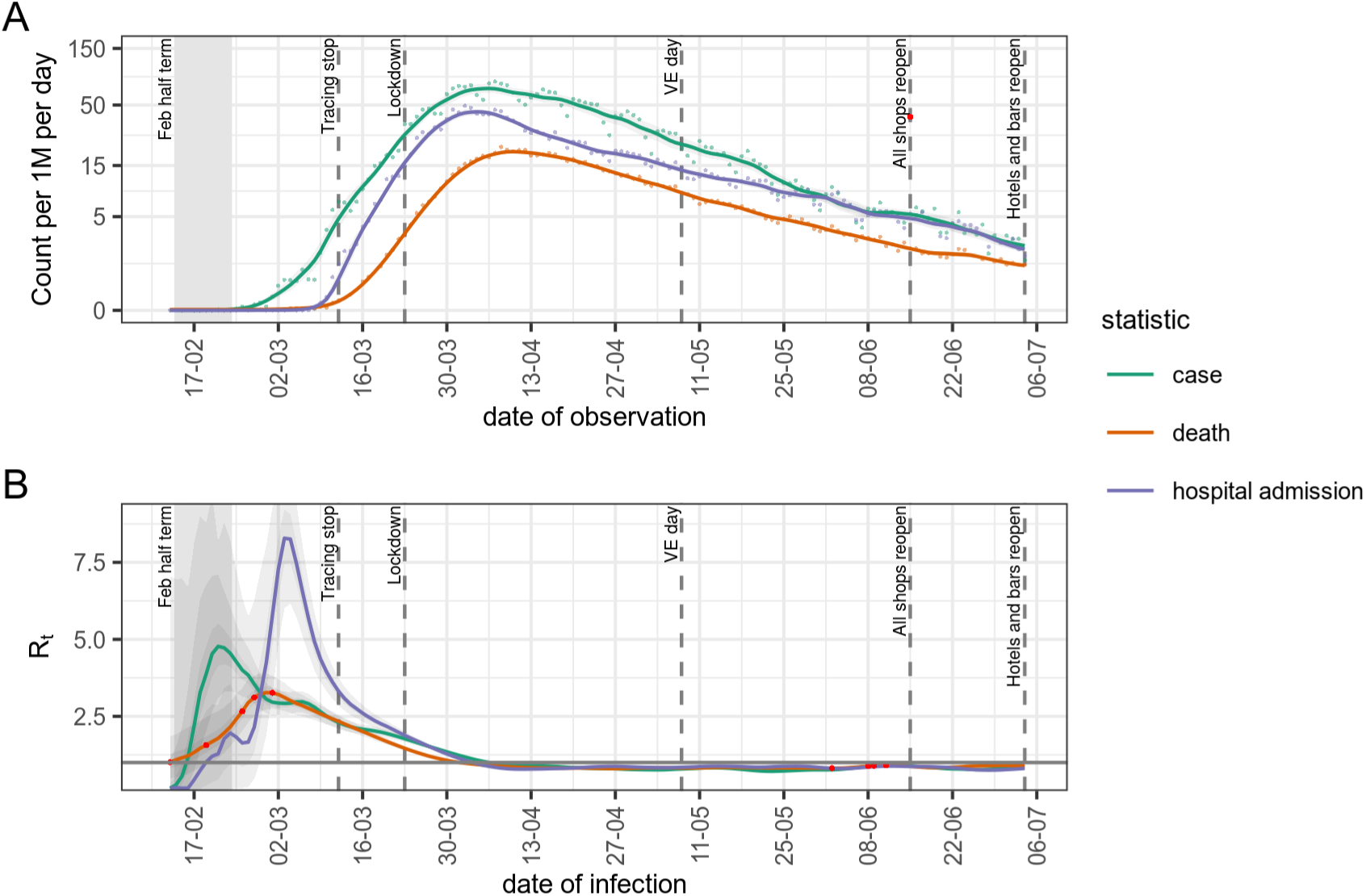
Timeline of cases and estimates of *R*_*t*_ based on cases reported by PHE and NHS labs (green), deaths reported in NHS trusts (red) and best available data for hospital admissions (blue). A) Number of cases (Pillar 1), deaths and admissions per million. B) estimates of *R*_*t*_ Red points are either missing values or identified as anomalies and replacements imputed.

### Countries in the UK

Estimates of *R*_*t*_ in the different countries of the UK, based on cases, deaths, hospital admissions or triage calls for the 4th July 2020 are presented in Table 2 and Figure 2. Triage figures based on 111 & 999 coronavirus pathway calls were available for England only; we do not have access to the full time series of all information for all countries. We show estimates based on a 28 day rolling window as cases and deaths in Northern Ireland, and Scotland, have fallen to a level which makes estimates over a shorter window unreliable. With insufficient information the Bayesian method used reverts to the prior value of *R*_0_ supplied which we set to 1. These estimates show the median value of *R*_*t*_ is below 1 for all four nations, using all data sources. The pattern in all nations is similar with *R*_*t*_ rapidly decreasing following lock-down on the 23rd March and becoming less than one in early April. Northern Ireland and Scotland have maintained a lower *R*_*t*_ for a longer period of time than England and Wales, with *R*_*t*_ values in Northern Ireland and Scotland at or below 0.75 for much of May, June and July compared to those in England and Wales which have been between 0.75 and 1 for the same period.

**Figure 2:**
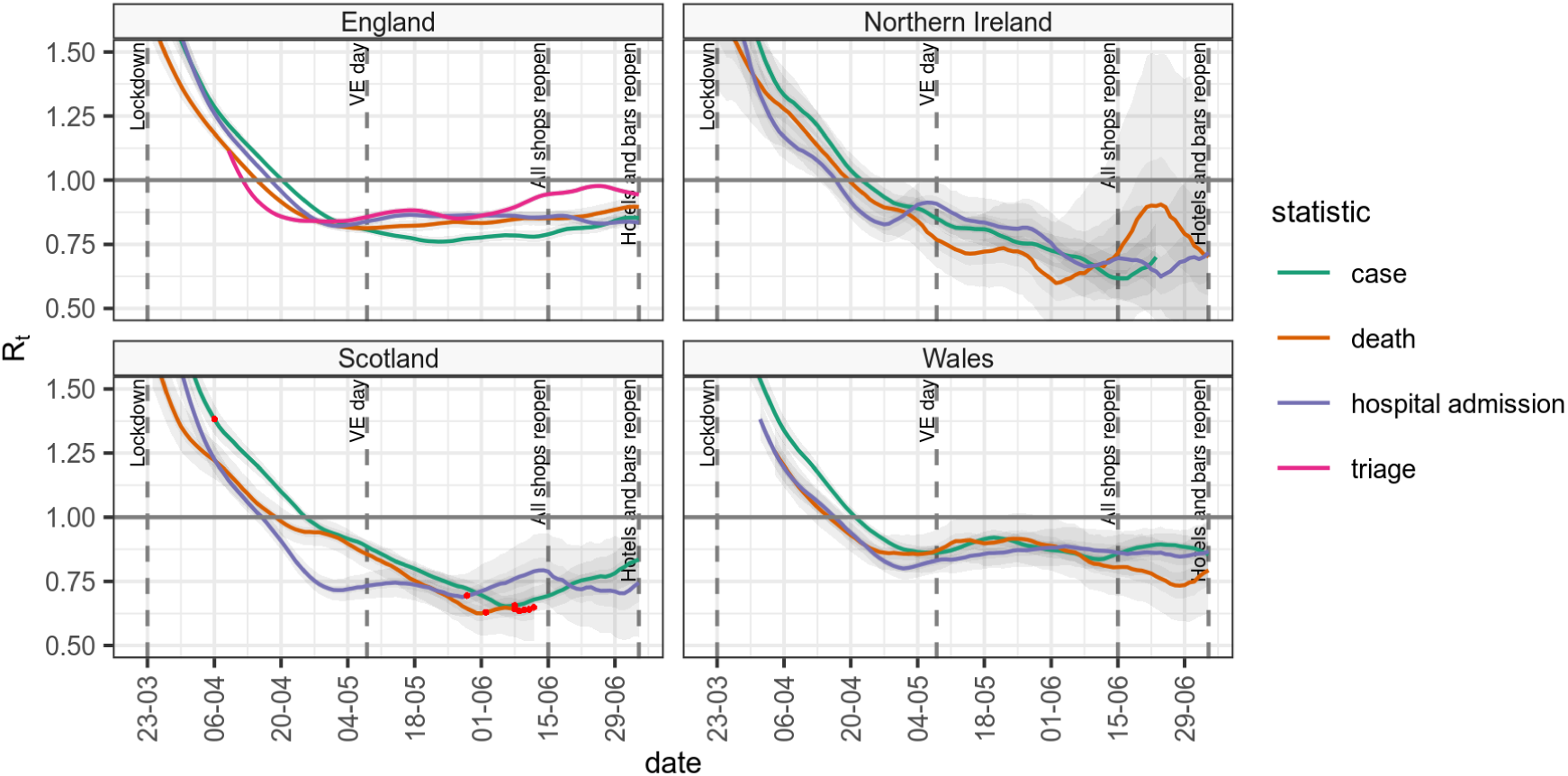
The median value of *R*_*t*_ and 95% confidence intervals for the individual countries in the United Kingdom, based on cases, deaths and hospital admissions, and a 28 day rolling window. For each data source, red points represent data points that were missing and have been imputed

### *R*_*t*_ by England NHS region

In Figure 3 (and, for completeness, Supplemental table 3) we present the same results as above but with a focus on the NHS regions in England. On the 4th July 2020 the estimates of the mean of *R*_*t*_ were largely below 1 in the individual NHS regions. The observed count of different observations demonstrates a clear regional difference between London and the South West and the rest of the country with lower rates of all indicators than other regions. The time series of regional estimates of *R*_*t*_ reflect the overall patterns of England, albeit with more volatility due to the different windowing size between Figure 2 and Figure 3. It is however possible to see higher levels of uncertainty in the South West reflecting the smaller case numbers and to a lesser extent the same in London.

**Table 3:** Estimates of the burden of disease before and after lock-down in the different NHS regions.

| NHS Region | Cases per 1M per day (95% CI) on 21 Mar | Cases per 1M per day (95% CI) on 04 Jul |
| --- | --- | --- |
| London | 49.6 (45.0; 54.6) | 1.7 (1.3; 2.1) |
| South East | 17.4 (16.0; 19.1) | 2.5 (2.0; 3.3) |
| South West | 7.8 (6.6; 9.1) | 0.4 (0.3; 0.6) |
| East of England | 14.7 (12.4; 17.4) | 2.7 (2.1; 3.6) |
| Midlands | 20.8 (18.9; 23.0) | 2.3 (1.8; 3.0) |
| North East and Yorkshire | 13.5 (12.3; 14.8) | 2.4 (2.1; 2.8) |
| North West | 16.0 (14.3; 17.9) | 2.6 (2.1; 3.2) |

**Figure 3:**
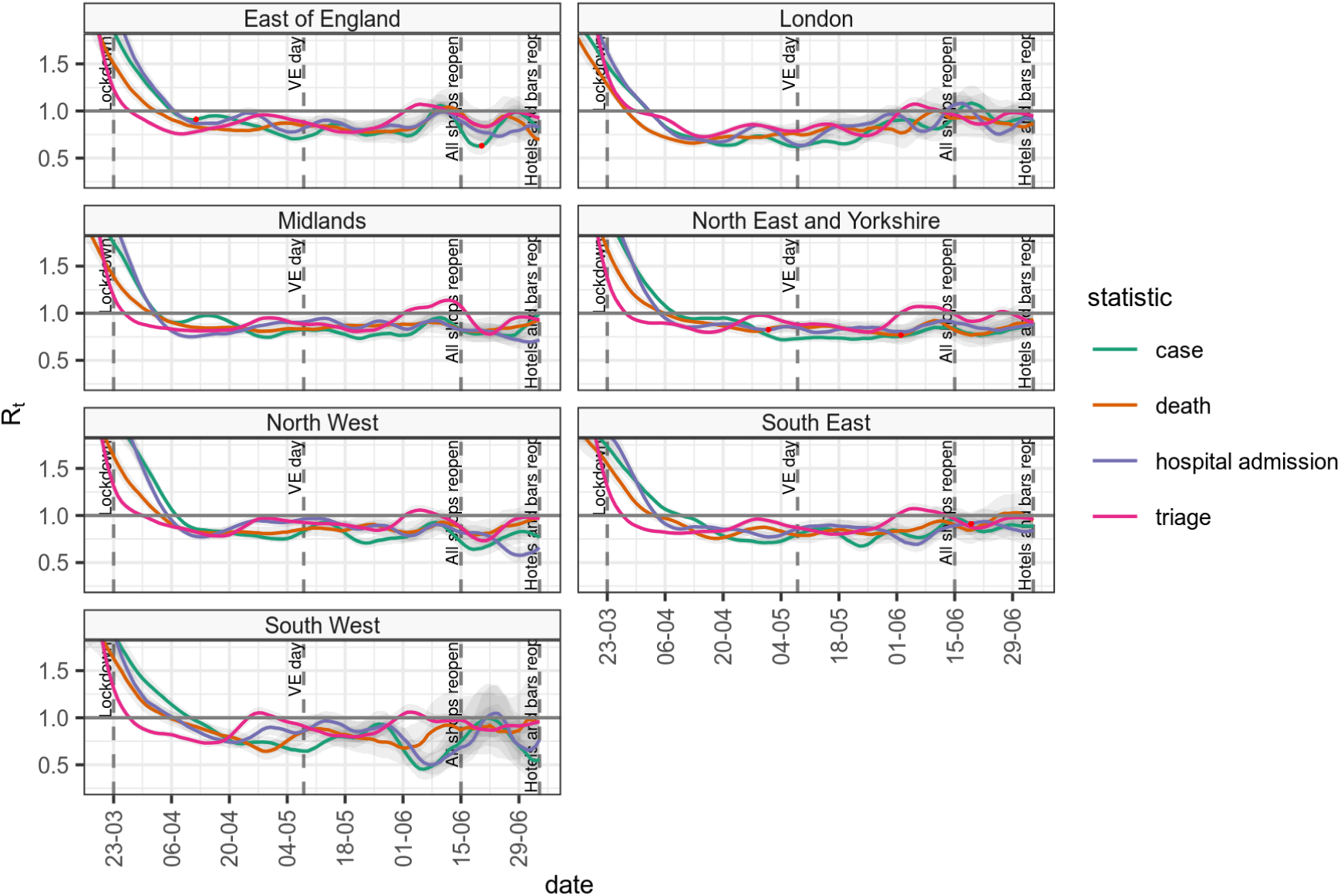
The median value of *R*_*t*_ and 95% confidence intervals for the sub-national regions of NHS England, based on cases, deaths and hospital admissions, and a 7 day rolling window.

### *R*_*t*_ differences from England baseline

In Figure 4 we plot the absolute difference of *R*_*t*_ in the seven NHS England administrative regions, from the *R*_*t*_ of England overall, as a baseline for each data source. This is based on 28 day window estimates due to the volatility observed in Figure 3. This analysis highlights the regional differences in *R*_*t*_ over time. Over the period of the lock-down, the East of England, Midlands and South East regions approximately tracked the England baseline. The South West was seen to initially following the national average but from June demonstrates a trend towards a lower value, although our confidence in these estimates is low. When lock-down started, London was initially well below the England average, by most indicators, and this continued until some point in late May, after which the trend reversed. On the other hand the North West, and less so North East & Yorkshire were consistently above the rest of the country until late May when they eventually reached the England average, and since mid June the *R*_*t*_ estimates in the North West have been lower than this average. The impact this variation has had on case loads is seen in Table 3 which shows the pre and post lock-down rates of newly identified cases in each NHS region.

**Figure 4:**
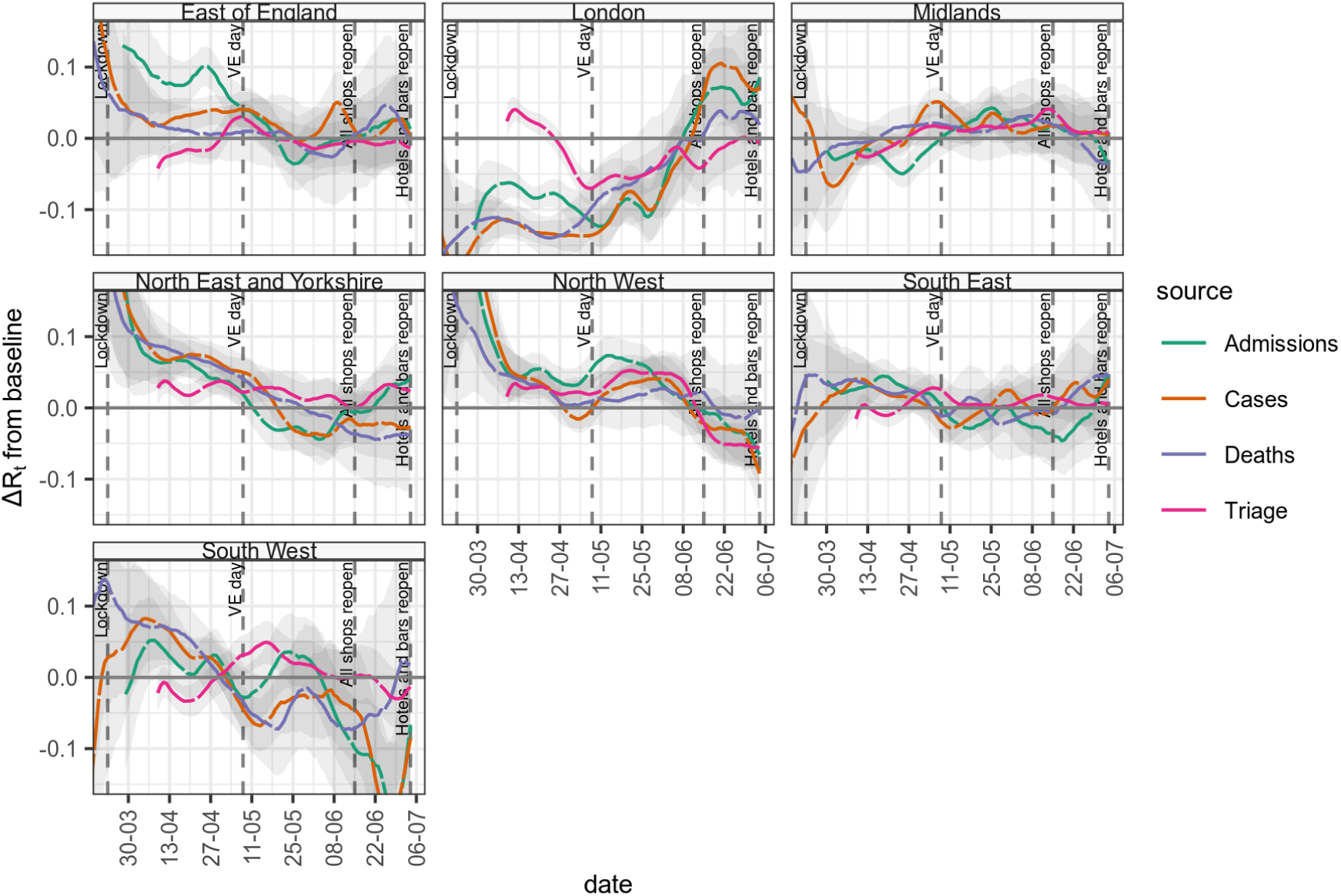
The difference of *R*_*t*_ estimates for NHS regions and baseline *R*_*t*_ estimates for England, based on cases, deaths and hospital admissions, and a 28 day rolling window.

## Discussion

In this paper, we estimated the reproduction number, *R*_*t*_, for COVID-19 in the UK using multiple data sources over a range of geographies and time points, and the same methodology. We find that, after the initial peak, different data sources produce similar results, without evidence of systematic bias for estimates based on more immediate measures (cases), compared to those based on longer term measures (deaths). In the UK, *R*_*t*_ peaked in mid February 2020, coinciding with the end of the school holidays. We find that using different data sources affects the estimated size of peak burden although with considerable uncertainty.

We find that *R*_*t*_ declined rapidly following lock-down on the 23rd March but did not reach the critical threshold (*R*_*t*_ less than one) separating growth and decline until early April. This delay is likely to be the combination of ongoing transmission within households, care homes and hospitals, or outbreaks in factories in key industries such as food production. Other factors that may influence the timing are delays in case identification and reporting and limitations in the estimation methods. Subsequent to mid-April we estimate Rt largely remained below one until the end of the lock-down period.

The multiple data sources we considered each have their advantages and drawbacks. Counts of test-positive cases and telephone triage calls provide a rapid indication of infection risk and capture a broad representation of age groups, but may be influenced by changes in behaviour and testing policy. In the UK, initial attempts at community tracing were abandoned when case numbers started to outstrip test availability and afterwards testing was only performed on hospital admissions for suspected COVID-19 [6]. Later, test capacity was increased and the policy reversed to include more community cases, again altering the nature of the population being tested. Although a regional breakdown of testing capacity was not available at the time of this analysis, we do know that capacity was exceeded in the early phase of the epidemic, and this is one reason why case based Rt estimates must be interpreted with caution until the middle of April. Hospital admissions and death data are less subject to changes in sampling strategy, although are subject to reporting delays and biases in ascertainment. As COVID-19 mortality is overwhelmingly in the elderly, statistics based on deaths mainly represent older groups. Due to reduced contact in the elderly we propose the outbreak took longer to become established in those age groups. Counts of admissions may be unreliable when there is delay in identifying a COVID-19 case, or when there is significant hospital transmission, as was the case in the early outbreak.

We took a pragmatic approach for sub-national analyses, based on data availability. In early April 2020, immediately following lock-down, estimates of *R*_*t*_ were lowest in London and highest in the North West and North East of England. At this time, the burden of cases was highest in London at nearly 50 cases per million people per day, but this reduced 25 fold over the lock-down period. In early July 2020, at the relaxation of lock-down, the case rates per capita were lowest in the South West, due to the smaller initial outbreak size, followed by London due to the larger impact of social distancing. However the benefit of lower case rates in London were offset by relative increases in *R*_*t*_. When case numbers are low *R*_*t*_ ceases to be a uniform statistic over a geographical area, because significant town to town variation will exist as clusters of infection become apparent. This was seen in South West of England towards the end of the lock-down where increasing variability in regional estimates of *R*_*t*_ became less obviously significant. This reinforces the point that *R*_*t*_ is a relative measure and should not be interpreted without information about the incidence rate.

Our approach has a number of limitations. Our method treats the whole system and each region as independent and isolated. In reality, regions are connected via travel, although this was reduced during early social distancing measures in March and April. Furthermore, in the early phases, importation rates were high [41], and this would lead to our approach overestimating the true *R*_*t*_.

## Conclusion

We present a description of the methodology and data sources used in providing estimates of *R*_*t*_ in the UK for SPI-M [14]. Our approach is pragmatic and designed to produce timely, useful information to policy makers. Despite this, we find that using a number of data sources and careful interpretation helps elucidate the regional differences in *R*_*t*_ and shows they existed from the outset of lock-down and persisted during lock-down. Due to compound nature of *R*_*t*_ the result of this variation is higher case loads in Northern regions in the UK exiting lock-down.

As we move forward early detection and prevention of spread of emerging clusters SARS-CoV-2 infections is critical to prevent large scale outbreaks. This will be challenging as the long incubation period and high rate of asymptomatic individuals makes undetected rapid spread easy. Prediction at a more localised level is needed to focus both community testing and more targeted social interventions on high risk areas in the future.

Critical to this work continues to be rapid access to information about the spread of SARS-CoV-2 in the community both with high spatial resolution, but also with a short time lag from infection to observation. As test and trace activities ramp up we expect to see similar biases in case related data as testing volumes change locally in response to outbreaks. Our existing approaches to estimating *R*_*t*_ principally use hospital based metrics and as such may not provide the perspective on the outbreak that is needed in the future. Telephone triage data is one potential source of information about local outbreaks, and an area for future investigation [42].

## Data Availability

All data is public domain and accessible through GitHub

https://github.com/terminological/uk-covid-datatools/vignettes/current-rt

## Supplementary material

**Supplemental table 1:** The combined data sources

| Resolution | Measure | Data source | Processing |
| --- | --- | --- | --- |
| CTRY | case | 4 nations case counts from 4 national PH sites, retrieved as timeseries from Tom White's github site, England pillar 1 tests from linelist | Cumulative case counts are converted to incidence figures. Pillar 1 only tests preferred in England, over PH data |
|  | death | SPI-M data feeds considered:<br>chess_death_inc_line,<br>dashboard_daily_death_inc_dash,<br>ons_death_inc_line,<br>nisra_death_inc_line,<br>sitrep_death_inc_line,<br>phw_death_inc_line | Feeds selected: chess_death_inc_line, sitrep_death_inc_line, phw_death_inc_line; combined by average if more than one per country |
|  | hospital admission | Various SPI-M data feeds: hospital_inc, hospital_inc_acuteone, hospital_inc_new, hospital_inc_new_acuteone, datereport_hospital_inc_new, spim_hosp_inc_new, spim_hosp_inc_new_authorisation_date | Hospital_inc_new & Hospital_inc_new_acuteone data set excluded for Wales. Hospital_inc_new and hospital_inc fields combined by summation, result and other sources combined by average, excluding missing values |
|  | triage | SPI-M 111 & 999 feed (only available for England) | Selected only calls with outcomes with 1 or 2 hour urgent clinical review, or ambulance dispatch. Daily 111 & 999 case counts added together. |
| NHSE | case | SPI-M line list | Line list aggregated by age and gender. Pillar 1 tests only as testing volumes variable in Pillar 2. |
|  | death | SPI-M deaths line list | Various care settings aggregated by summation. Result is all deaths in all locations |
|  | hospital admission | SPI-M hospital_inc and hospital_inc_new fields | Sources combined by summation |
|  | icu admission | SPI-M chess_icu_admissions field | none |
|  | triage | SPI-M 111 feed | Selected only calls with outcomes with 1 or 2 hour urgent clinical review, or ambulance dispatch. |
| UK | case | UK headline from PHE and 4 nations case counts from 4 national PH sites, retrieved as timeseries from Tom White's github site for Wales, Scotland and Northern Ireland. Pillar 1 tests from line list for England. | Ultimately sum of 4 nations case counts previously selected for CTRY cases used as headline numbers variably include Pillar 2. Cumulative case counts are converted to incidence figures. |
|  | death | Sum of 4 nations figures previously selected for CTRY deaths | none |
|  | hospital admission | Sum of 4 nations figures previously selected for CTRY hospital admissions | none |

**Supplemental table 2:** Estimated correction of *R*_*t*_ offset based on incubation period and observation delay

| Observation | Mean delay (days) | SD (days) | 95% quantiles (days) |
| --- | --- | --- | --- |
| onset | 5.42 | 5.89 | 0.64; 20.86 |
| test | 7.15 | 6.98 | 0.50; 24.74 |
| admission | 8.03 | 10.03 | 0.72; 35.53 |
| death | 17.46 | 13.31 | 3.47; 54.13 |

**Supplemental table 3:** Estimates of mean values of *R*_*t*_ and 95% confidence intervals for the sub-national regions of NHS England, based on 111 calls, cases, deaths and hospital admissions on the 4th July 2020

| NHS Region | Observation | R(t) (95% CI) | Count per 1M per day (95% CI) |
| --- | --- | --- | --- |
| East of England | Cases | 0.81 (0.67; 0.97) | 2.7 (2.1; 3.6) |
|  | Deaths | 0.70 (0.53; 0.91) | 1.6 (1.4; 1.9) |
|  | Admissions | 0.83 (0.68; 0.99) | 2.6 (2.0; 3.3) |
|  | Triage | 0.92 (0.86; 1.00) | 12.1 (10.5; 13.9) |
| London | Cases | 0.90 (0.74; 1.08) | 1.7 (1.3; 2.1) |
|  | Deaths | 0.87 (0.64; 1.14) | 0.9 (0.8; 1.2) |
|  | Admissions | 0.91 (0.79; 1.05) | 2.7 (2.1; 3.6) |
|  | Triage | 0.94 (0.87; 1.02) | 8.4 (7.4; 9.6) |
| Midlands | Cases | 1.00 (0.87; 1.14) | 2.3 (1.8; 3.0) |
|  | Deaths | 0.93 (0.77; 1.11) | 1.8 (1.6; 2.1) |
|  | Admissions | 0.72 (0.61; 0.84) | 2.5 (2.2; 2.9) |
|  | Triage | 0.93 (0.88; 0.98) | 15.3 (13.9; 16.8) |
| North East and Yorkshire | Cases | 0.88 (0.75; 1.03) | 2.4 (2.1; 2.8) |
|  | Deaths | 0.94 (0.76; 1.15) | 1.7 (1.4; 2.0) |
|  | Admissions | 0.88 (0.76; 1.01) | 2.9 (2.6; 3.4) |
|  | Triage | 0.90 (0.85; 0.95) | 17.1 (15.2; 19.2) |
| North West | Cases | 0.77 (0.64; 0.92) | 2.6 (2.1; 3.2) |
|  | Deaths | 0.98 (0.81; 1.18) | 2.1 (1.9; 2.4) |
|  | Admissions | 0.66 (0.55; 0.78) | 3.0 (2.6; 3.4) |
|  | Triage | 0.97 (0.90; 1.04) | 13.5 (11.3; 16.2) |
| South East | Cases | 0.89 (0.76; 1.03) | 2.5 (2.0; 3.3) |
|  | Deaths | 0.95 (0.78; 1.14) | 1.5 (1.4; 1.7) |
|  | Admissions | 0.84 (0.70; 0.99) | 2.3 (1.9; 2.6) |
|  | Triage | 0.96 (0.90; 1.02) | 14.5 (12.9; 16.2) |
| South West | Cases | 0.56 (0.31; 0.92) | 0.4 (0.3; 0.6) |
|  | Deaths | 0.96 (0.63; 1.37) | 0.7 (0.5; 1.0) |
|  | Admissions | 0.76 (0.52; 1.09) | 0.7 (0.5; 0.9) |
|  | Triage | 0.96 (0.87; 1.05) | 9.0 (7.8; 10.5) |

